# Extended use or re-use of single-use surgical masks and filtering facepiece respirators: A rapid evidence review

**DOI:** 10.1101/2020.06.04.20121947

**Authors:** E Toomey, Y Conway, C Burton, S Smith, M Smalle, XH Chan, A Adisesh, S Tanveer, L Ross, I Thomson, D Devane, T Greenhalgh

## Abstract

**Background:** The COVID-19 pandemic has led to unprecedented demand for personal protective equipment. Shortages of surgical masks and filtering facepiece respirators has led to the extended use or re-use of single-use respirators and surgical masks by frontline healthcare workers. The evidence base underpinning such practices has been questioned.

**Objectives:** To summarise guidance and synthesise systematic review evidence on extended use, re-use or reprocessing of single-use surgical masks or filtering facepiece respirators.

**Methods:** A targeted search of the World Health Organization, European Centre for Disease Prevention and Control, the US Centers for Disease Control and Prevention, and Public Health England websites was conducted to identify guidance. Four databases (Medline, Pubmed, Epistemonikos, Cochrane Database of Systematic Reviews) and three preprint repositories (Litcovid, MedRxiv and Open Science Framework) were searched for relevant systematic reviews. Record screening and data extraction was conducted by two reviewers. Quality of included systematic reviews was appraised using the AMSTAR-2 checklist. Findings were integrated and narratively synthesised to highlight the extent to which key claims in guidance documents were supported by research evidence.

**Results:** Six guidance documents were identified. All note that extended use or re-use of single-use surgical masks and respirators (with or without reprocessing) should be considered only in situations of critical shortage. Extended use was generally favoured over re-use because of reduced risk of contact transmission. Four high-quality systematic reviews were included: three focused on reprocessing (decontamination) of N95 respirators and one focused on reprocessing of surgical masks. There was limited evidence on the impact of extended use on masks and respirators. Vaporised hydrogen peroxide and ultraviolet germicidal irradiation were highlighted as the most promising reprocessing methods, but evidence on the relative efficacy and safety of different methods was limited. We found no well-established methods for reprocessing respirators at scale.

**Conclusions:** There is limited evidence on the impact of extended use and re-use of surgical masks and respirators. Where extended use or re-use is being practiced, healthcare organisations should ensure that policies and systems are in place to ensure these practices are carried out safely and in line with available guidance.

## Background

The COVID-19 pandemic has put healthcare systems under unprecedented strain and exposed healthcare workers in a wide range of clinical environments to risk of serious infection^1^. Infection prevention and control measures developed for healthcare workers recommend personal protective equipment including surgical masks and respirators as one part of a broader hierarchy of protective measures^2^. Surgical masks protect the mouth, nose, and lower face against splashes and droplets and have some limited but variable filtration ability and do not seal effectively to the face. Therefore, they are not regarded as protective against inhalation of small (< 5μm) aerosolised particles including airborne pathogens. Filtering facepiece respirators (including N95, FFP2 and FFP3 specifications) are single-use masks which protect against inhalation of aerosolised particles. They do this by fitting tightly to the face to ensure that inhaled air passes through a filter layer within the mask; fit-testing is essential.

Global shortages have forced protective equipment-sparing measures, including extended use, re-use and consideration of the reprocessing of single-use masks and respirators^3^. Extended use is the practice of using the same single-use mask or respirator for encounters with multiple patients, without removing it^4^. Re-use is the practice of using the same mask or respirator for multiple encounters with patients, removing it (‘doffing’) for storage after each encounter and putting it on again (‘donning’) prior to the next encounter with a patient^4^. Reprocessing is ‘decontamination using disinfection or sterilization methods followed by reuse of either reusable or disposable PPE’^5^. When applied to single-use masks and respirators, each of these practices can potentially lead to reduced respiratory protection, comfort and safety for healthcare workers (Figure 1).

**Figure 1.**
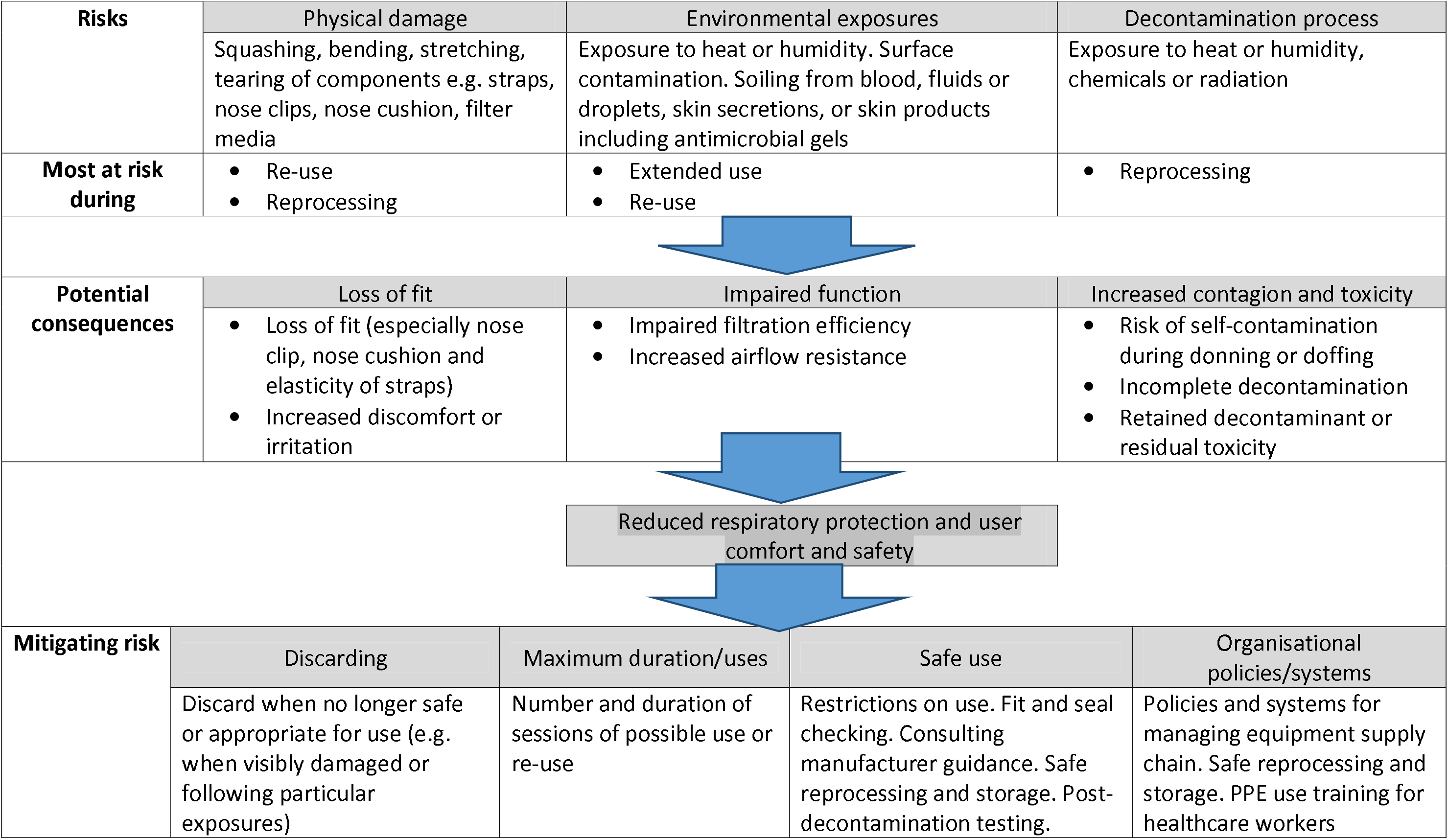
Taxonomy of potential risks and mitigation with respect to extended use/re-use/reprocessing of single-use masks and respirators

A number of national and international guidelines make reference to extended use, re-use and reprocessing of single-use masks and respirators^6^. We compared these guidelines first with each other and second with current synthesised evidence, particularly in the light of current worldwide shortages of personal protective equipment, in order to inform rapidly evolving policies and practice.

## Methods

We conducted a rapid review in line with the Cochrane Interim Guidance for Rapid Reviews^7^. The project workflow and initial protocol were published on Open Science Framework^8^ on April 6^th^ 2020 with a final revised protocol published on May 5^th^ 2020 before completion of data extraction. This review is reported according to the PRISMA reporting criteria for systematic reviews (Appendix 1)^9^. It is one of a suite of reviews on personal protective equipment undertaken as part of the Oxford COVID-19 evidence series and edited by TG.

### Searches and identifying literature

#### Identification of guidance

ET and MS carried out a targeted search of major international and national health organisation websites including the World Health Organization, European Centre for Disease Prevention and Control, the US Centers for Disease Control and Prevention, and Public Health England between March 23^rd^ and May 22^nd^2020 to identify current recommendations and documents providing specific guidance on re-use or extended use of surgical masks or filtering facepiece respirators. Identified documents were screened for inclusion by ET and verified by YC.

#### Identification of systematic reviews

We aimed to identify systematic reviews of primary studies exploring any aspect of the extended use, reuse or reprocessing of any type of surgical mask or filtering facepiece respirator on outcomes including technical performance standards, decontamination outcomes, or impact on healthcare workers (e.g. health outcomes or qualitative outcomes such as acceptability). Systematic electronic database searches of Medline, the Cochrane Database of Systematic Reviews, Epistemonikos and PubMed were conducted by MS, an experienced information specialist, on April 28^th^ 2020. This was supplemented with searches of preprint repositories Litcovid, MedRxiv, and OSF as well as using Google Scholar as a control check. We scanned reference lists of included documents for additional relevant records and contacted authors of included reviews. No date limit or language restrictions were applied. Search terms included (masks OR respiratory devices) AND (infection control OR decontamination) AND (re-use OR extended use). A systematic review search filter was not applied in Medline, but was applied in Epistemonikos, Cochrane and PubMed to minimise duplication and increase specificity. A complete search strategy is provided in Appendix 2. Records were imported into Covidence and all records were double-screened by two reviewers (ET, ST) independently at both title/abstract and full text stage.

### Data extraction and synthesis

Data were extracted by ET and verified by YC using separate pre-developed data extraction templates in Excel and NVIVO for the two sections of this review. Data extraction fields included categories (e.g. organisation, country) and free text (e.g. definitions of terms, details of recommendations, review findings). Data extracted from guidelines were tabulated according to recommendations regarding extended use, reuse and reprocessing for surgical masks and filtering facepiece respirators. No quality appraisal was undertaken on guidelines since the purpose of this part of the review was to document what was being recommended. Included systematic reviews were assessed independently for quality using the using the AMSTAR 2 checklist for applied by experienced systematic reviewers (DD, ET). Data extracted from systematic reviews were tabulated to compare across reviews and integrate with findings from the review of standards.

Findings from the data extraction were integrated by ET and CB using a narrative synthesis approach with critical review and appraisal from YC, who checked to ensure that narrative synthesis reflected the original findings of source documents. Critical input was also provided by topic experts in infection control (XHC, LR), occupational medicine (AA), and personal protective equipment (SS) to ensure relevance and applicability.

## Results

### Search results

Six documents (from four organisations) were identified that provided guidance on the extended use, reuse and/or reprocessing of surgical masks or filtering facepiece respirators (overview provided in Appendix 3). The search for reviews retrieved 458 records. After duplicate removal and title and abstract screening, 60 full-text articles were screened. Four relevant systematic reviews were identified and included in our review (Figure 2).

**Figure 2.**
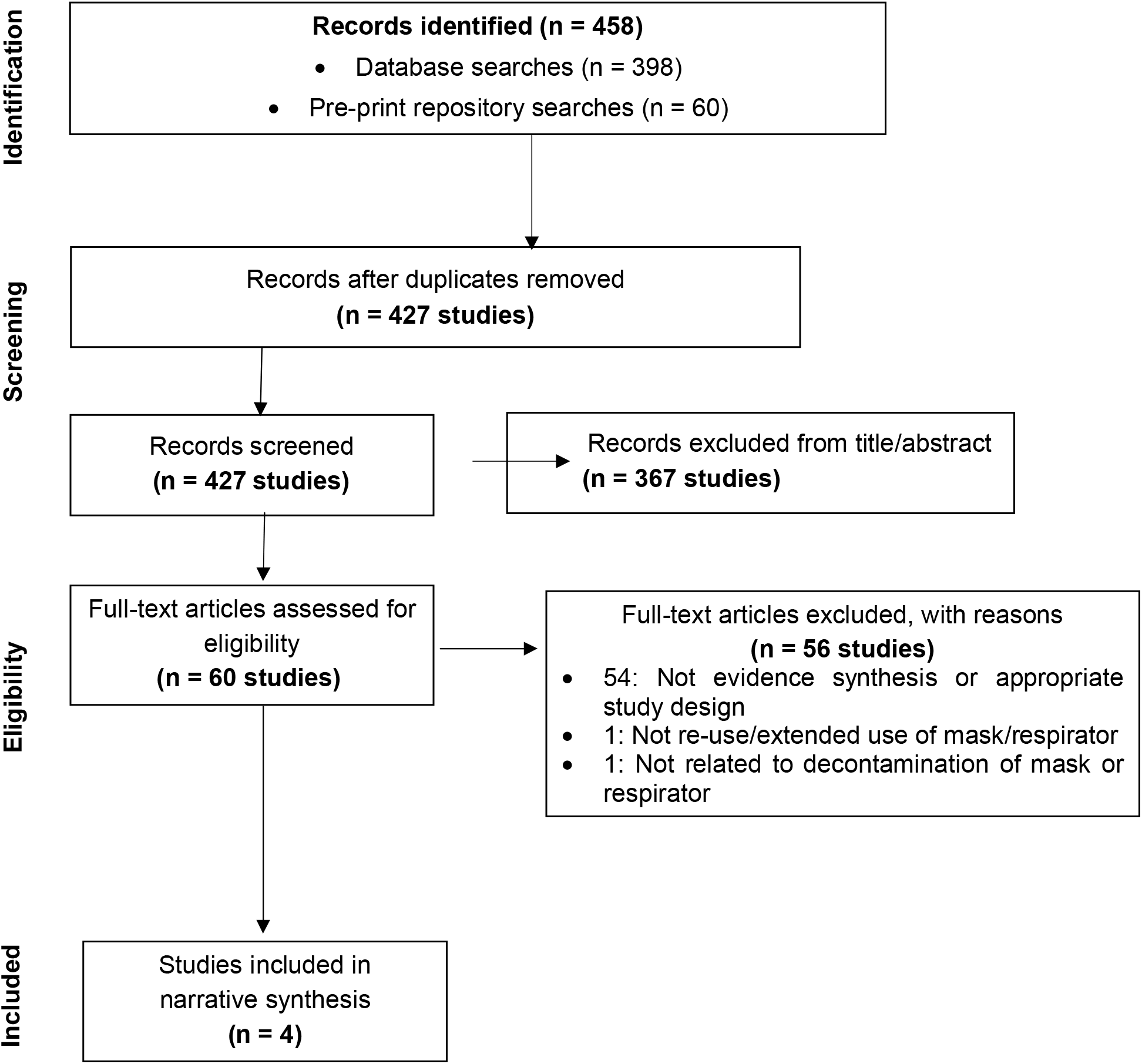
PRISMA flow diagram

### Summary of guidance documents

Three guidance documents were from the US Centers for Disease Control and Prevention^4 10 11^ and one each from the European Centre for Disease Prevention and Control^12^, Public Health England^13^, and the World Health Organization^5^. All were published or updated between March 17^th^ 2020 and May 21^st^ 2020 and written for the COVID-19 pandemic. None of the guidance documents included a systematic literature search and the depth of referencing varied. US Centers for Disease Control and Prevention guidance was the most thorough in terms of citing evidence to inform recommendations.

The scope of the guidance documents varied. US Centers for Disease Control and Prevention guidance covered extended use, re-use and reprocessing. The European Centre for Disease Prevention and Control guidance covered only reprocessing. Public Health England covered extended use and re-use. World Health Organization guidance covered extended use and reprocessing. All four guidance documents referred to respirators but varied in their coverage of surgical masks. The US Centers for Disease Control and Prevention and World Health Organization guidance discussed surgical masks and respirators separately. Public Health England did not differentiate between the two. The European Centre for Disease Prevention and Control made limited reference to surgical masks.

All guidance documents depicted extended use, re-use or reprocessing of single-use masks and respirators as extraordinary, last-resort measures to be considered only during critical shortage of equipment and when other strategies for rational use and conservation of supplies have been exhausted (e.g. minimising need for PPE through administrative and engineering controls, coordinating supply chain management). The US Centers for Disease Control and Prevention, World Health Organization, and Public Health England recommend additional procedures at organisational level such as appropriate documentation and recording of re-use or reprocessing, quality assurance of reprocessing measures, suitable reprocessing and storage facilities and systems, and staff training regarding safe use and donning or doffing of masks or respirators if re-use or extended use.

All guidance documents favoured extended use over re-use because of reduced risk of contact transmission. All recommended ensuring good fit, performing a seal check, and inspecting for function and potential damage prior to use or re-use of any mask or respirator. The US Centers for Disease Control and Prevention and World Health Organization, acknowledged that facemasks and respirators that have passed their expiry date may sometimes be used in situations of limited capacity.

Figure 3 provides a schematic summary of current international guidelines on re-use, reprocessing, and extended use of single-use masks and respirators.

**Figure 3.**
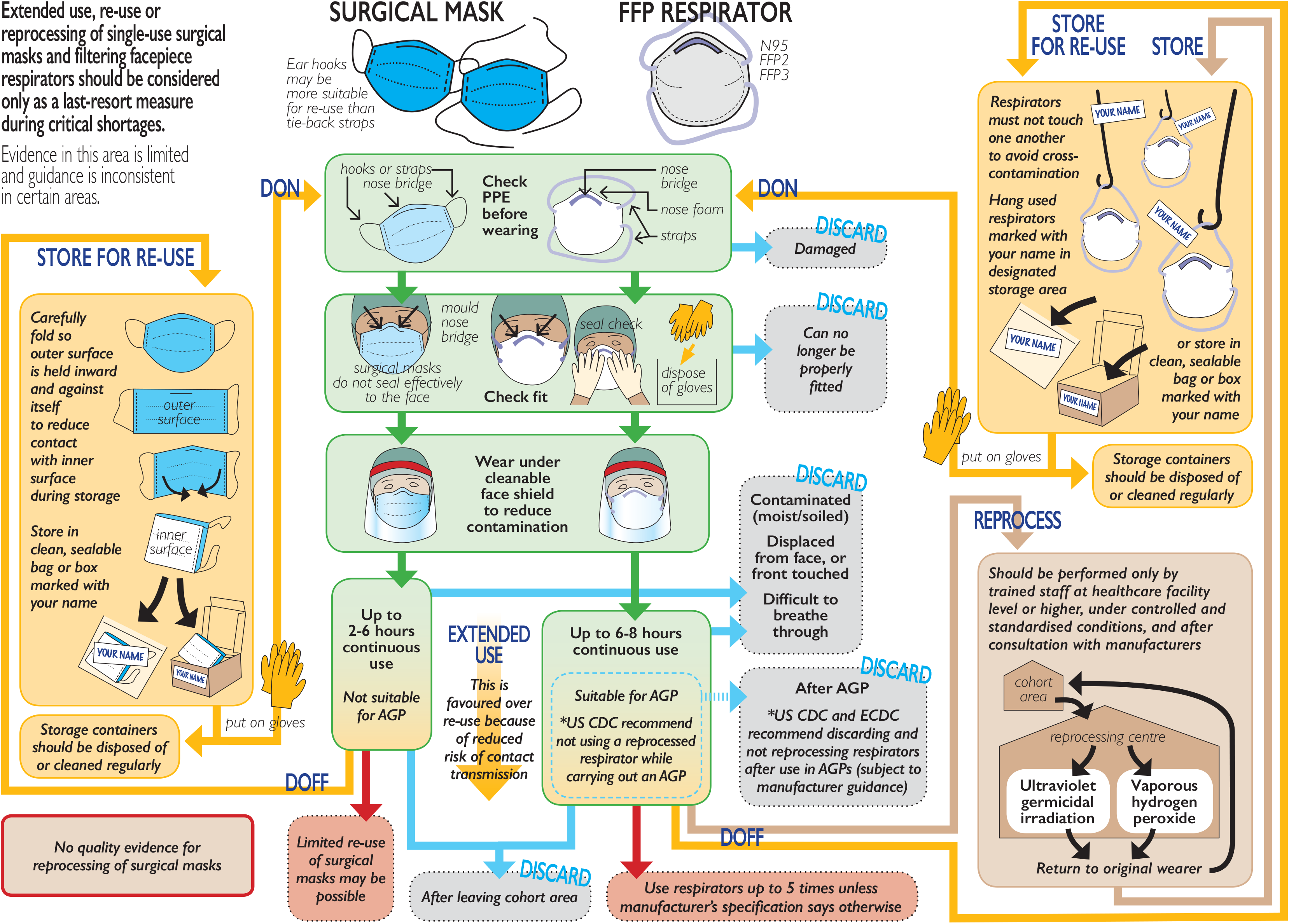

#### Surgical masks

Table 1 summarises guidance on extended use and re-use of surgical masks with findings grouped by the three forms of risk mitigation shown in Figure 1. The three sources of guidance are in broad agreement that extended use of surgical masks may continue while in a cohorted area (e.g. on a COVID-19 ward), but they differ on the time recommended (from 2 to 6 hours). There was substantial disagreement among these guidance documents on whether surgical masks may be re-used. None of the guidance documents recommended reprocessing of surgical masks.

**Table 1.** Summary of guidance recommendations for fluid resistant surgical masks.

|  | CDC | ECDC | PHE<sup>1</sup> | WHO |
| --- | --- | --- | --- | --- |
| **Extended use** |  |  |  |  |
| Discarding | Soiled, damaged or hard to breathe through | No information | Moist, damaged, visibly soiled or difficult to breathe through. | Wet, soiled, splashed, damaged or difficult to breathe through.<br>If displaced from face or touched |
| Maximum duration | Not specified |  | 2-6 hours dependent on setting and activity | Up to 6hrs |
| Safe use | Hand hygiene if adjusted/ touched<br>Only continue within cohorted area. |  | Hand hygiene if adjusted/ touched<br>Discard after leaving cohorted area unless transferring a patient | Only continue within cohorted area - discard on leaving |
| **Re-use** |  |  |  |  |
| Discarding | Soiled, damaged or hard to breathe through | Not recommended | Moist, damaged, visibly soiled or difficult to breathe through. | No information |
| Maximum uses | Not specified |  | Not specified |  |
| Safe use | Do not re-use masks that fasten with ties (elastic ear hooks more suitable)<br>Fold<sup>2</sup> carefully and store in sealable paper bag or breathable container between uses. |  | Masks with elastic ear hooks more suitable for re-use<br>Fold<sup>2</sup> carefully and store in labelled sealable bag between uses<br>Some models cannot be re-used as they deform once being donned |  |
| **Reprocessing** |  |  |  |  |
| Discarding | Not recommended |  |  |  |
| Maximum duration / uses |  |  |  |  |
| Safe use |  |  |  |  |

<sup>1</sup> PHE guidelines on extended use and re-use do not clearly differentiate between surgical masks and filtering facepiece respirators
<sup>2</sup> "Fold" – this cannot apply to moulded cup-type face masks

#### Filtering facepiece respirators

Table 2 summarises guidance on extended use and re-use of respirators. The World Health Organization guidelines recommend use for up to six hours and US Centers for Disease Control guidance noted up to eight hours of use in specific studies, however, they emphasise that duration should be guided primarily by hygienic or practical issues (e.g. shift ending) and that the device should be discarded if it becomes soiled, damaged or difficult to breathe through. The European Centre for Disease Prevention and Control and US Centers for Disease Control recommend that the addition of a cleanable face shield over a respirator to prevent splashes or contact may help extend the life of respirators. Regarding maximum number of re-uses, the US Centers for Disease Control proposes up to five uses (donnings) unless stated otherwise by the manufacturer; there was no consensus across documents.

**Table 2.** Summary of guidance recommendations for filtering facepiece respirators.

|  | CDC | ECDC | PHE<sup>1</sup> | WHO |
| --- | --- | --- | --- | --- |
| **Extended use** |  |  |  |  |
| Discarding | Soiled (blood, secretions, body fluids), if obvious damage or difficult to breathe through<br>Discard after use in aerosol generating procedure. | Soiled, wet, can no longer be properly fitted or difficult to breathe through<br>Discard after use in aerosol generating procedure | Moist, damaged, visibly soiled or difficult to breathe through. | Wet, soiled, splashed, damaged or difficult to breathe through.<br>If displaced from face or touched |
| Maximum duration | Not specified (notes some studies up to 8hrs) - guided by hygienic/practical concerns | Time guided by hygienic/practical concerns<sup>2</sup> | Not specified<sup>3</sup> | Up to 6 hours |
| Safe use | Hand hygiene if adjusted or touched. Respirator can be covered with face shield ('strongly preferred') or surgical mask. Discard after leaving cohorted area. | Respirator can be covered with face shield or medical mask to extend use | Hand hygiene if adjusted or touched. Discard after leaving cohorted area unless transferring a patient | Only continue within cohorted area - discard after leaving |
| **Re-use<sup>4</sup>** |  |  |  |  |
| Discarding | Soiled (blood, secretions, body fluids), if obvious damage or difficult to breathe through<br>Discard after use in aerosol generating procedure. | No information | Moist, damaged, visibly soiled or difficult to breathe through. | Re-use without re-processing strongly discouraged |
| Maximum uses | Up to 5 times unless manufacturer states otherwise. |  | Not specified |  |
| Safe use | (a) Ensure safe storage and limit to named user<br>(b) Follow one day of use with 4 day "quarantine" period in labelled breathable sealed container before re-use<sup>5</sup> |  | Fold and store in labelled sealable bag between uses<br>Some models cannot be re-used as they deform once being donned |  |
| **Reprocessing<sup>4</sup>** |  |  |  |  |
| Discarding | If integrity of any component (e.g. straps, bridge) of the respirator is compromised, or if a successful user seal check cannot be performed. | Not specified | Currently no recommendations (but acknowledge work on validating methods in progress) | Check integrity and shape before reprocessing – discard if damaged/not suitable for re-use |
| Maximum duration / uses | Not specified | Not specified |  | "After a pre-defined number of uses" |
| Safe use | Not for use in aerosol generating procedures<sup>6</sup> | Quality checks of reprocessing necessary to ensure safety |  | After use, label and place in reprocessing container<br>Reprocessing performed by trained staff<br>Return to original wearer after reprocessing |

<sup>1</sup> PHE guidelines on extended use and re-use do not clearly differentiate between surgical masks and filtering facepiece respirators
<sup>2</sup> "...can be re-used for a limited time, unless there is a risk for contamination through the deposition of infectious particles on the surface"
<sup>3</sup> States that filter capacity of FFP3/FFP2/N95 respirators greatly exceeds one day of use in health/social care
<sup>4</sup> Re-used and reprocessed respirator are likely to be used for extended periods, in this case the extended use criteria also apply.
<sup>5</sup> Point (b) currently appears in decontamination guidelines as an alternative to decontamination, but does not appear in re-use guideline.
<sup>6</sup> unless manufacturer information indicates that decontamination does not affect performance

Table 2 also summarises guidance on reprocessing of respirators, which were framed with caution to reflect the high degree of uncertainty and potential risk. Notably, the US Centers for Disease Control and Prevention guidance, which provides the most detail regarding decontamination, states that reprocessed masks should not be used during aerosol generating procedures unless there is explicit guidance from the manufacturer that a respirator can be used after a particular form of reprocessing without negatively impacting its performance. Two documents emphasised the wide variability of respirator models for reprocessing, and the importance of consulting manufacturer guidance where possible. The World Health Organization guidance states that reprocessing should be performed by trained staff in the ‘sterile services department of a health care facility or at bigger scale under controlled and standardized conditions.’

### Systematic reviews

The four systematic reviews were conducted between March and April 2020 and at the time of writing were pre-prints or under peer-review. All were conducted in Canada, three by the same research team. Three focused on reprocessing of filtering facepiece respirators using different decontamination interventions (heat-based treatments, disinfectant, and ultraviolet germicidal irradiation),^14–16^ and one covered reprocessing of surgical masks and ‘pre-contamination’ interventions applied before use to enable extended use or re-use. No reviews in our sample examined the impact of extended use or re-use of filtering facepiece respirators or surgical masks on the ability to meet technical standards or on healthcare worker acceptability outcomes such as comfort.

AMSTAR 2 ratings for each study are provided in Appendix 4. Included systematic reviews were judged to be predominantly of high quality. However, none provided a list of excluded studies or justified the reasons for exclusion, and no study reported on the sources of funding for included primary studies.

Detailed descriptions of the reviews and key findings are provided in Appendix 5. The three reviews on filtering facepiece respirators included between 11 and 13 studies (28 unique studies in total). Primary studies included in these reviews had evaluated the effects of reprocessing on various outcomes including effective decontamination, appearance and performance (including filtration efficiency and airflow resistance), user comfort, fit and safety. In relation to decontamination methods, these reviews found most evidence to support vaporised hydrogen peroxide, moist/dry heat in the range 60–90^9^C, and ultraviolet germicidal irradiation interventions. Studies included in the reviews were generally at low risk of bias. However, few primary studies investigated the impact of these methods on fit or user comfort, and there was substantial variability in the models of filtering facepiece respirator used across studies.

Only two studies^17^ ^18^ included in one review^16^ explored the effect of reprocessing on SARS-CoV-2. Seven studies were included in the review of surgical mask decontamination, but only one of these had specifically evaluated decontamination interventions after use to enable re-use. The review concluded that there was insufficient evidence regarding the safety or efficacy of any decontamination intervention for reprocessing surgical masks.

### Comparison of guidance and systematic reviews on reprocessing of masks or respirators

Table 3 compares the findings of the systematic reviews with the three guidance documents relating to reprocessing of surgical masks and respirators. There is considerable discrepancy to the extent that no single reprocessing method is supported by all the guidance documents. The intervention with most support is vaporised hydrogen peroxide, though one document cautions about chemical residues and another indicates it has only been tested with some of the respirator models in common use. Similarly, ultraviolet germicidal irradiation receives both cautious support and concerns about inadequate decontamination because of incomplete penetration into deeper layers of the filter. Moist heat is cited as promising, though there are concerns when steam is microwave-generated where there may be uneven heating and where the metal nose band may generate sparks.

**Table 3.**
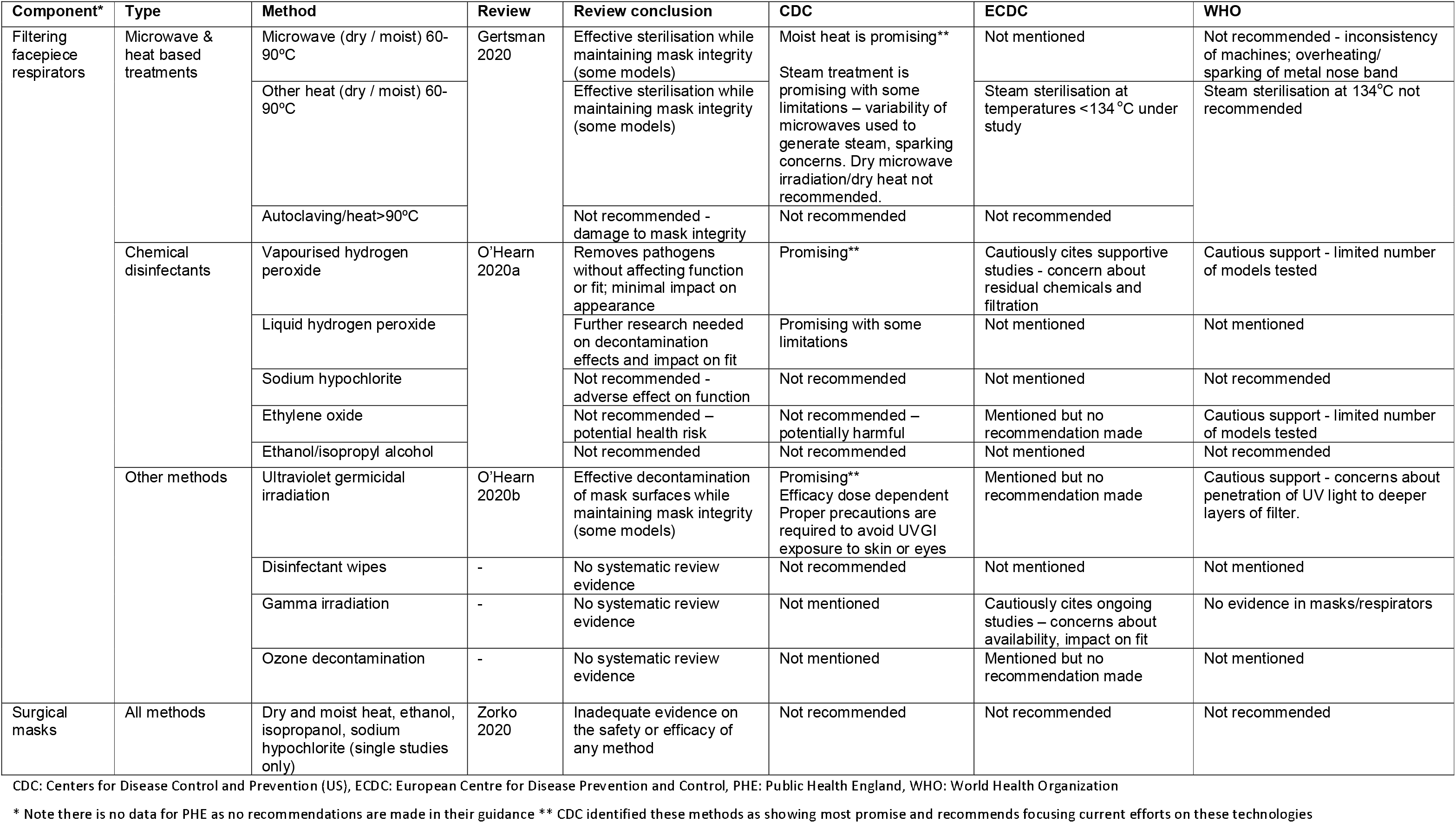
Summary of systematic review conclusions compared to guidance recommendations for reprocessing methods.

### Inconsistencies and gaps in the evidence

Our review of guidance documents and systematic reviews highlights several inconsistencies that warrant exploration, even taking account of the fact that the COVID-19 pandemic presents unique challenges for different contexts. This is particularly true of the emerging science of reprocessing of respirators, for which guidance authorities appear to have proceeded with different degrees of haste or caution in making recommendations.

Although the European Centre for Disease Prevention and Control recommend addition of either a surgical mask or a cleanable face shield over a respirator, the US Centers for Disease Control and Prevention strongly recommends a face shield rather than a surgical mask due to supply issues and concerns that a surgical mask could affect the function of the N95 respirator. Moreover, evidence suggests addition of a surgical mask does not improve respiratory protection and may increase user discomfort and impair communication^19 20^

The US Centers for Disease Control and Prevention and European Centre for Disease Prevention and Control guidance documents make specific recommendations in relation to respirators and aerosol generating procedures. Both recommend discarding and not reprocessing respirators after use in aerosol generating procedures, and the US Centers for Disease Control and Prevention also recommend not using a reprocessed respirator while carrying out an aerosol generating procedure (subject to manufacturer guidance). However, the World Health Organization and Public Health England do not explicitly recommend respirators be discarded after aerosol generating procedures, nor do they mention the use of reprocessed respirators in aerosol generating procedures. No guidance mentions the use or discarding of respirators in areas where aerosols may be present.

There is inconsistency and lack of clarity regarding the re-use of respirators without reprocessing. The World Health Organization guidance strongly discourages re-use of respirators without appropriate decontamination. However, citing a recent study by Van Doremalen et al. suggesting that SARS-CoV-2 viral particles may stay infective for up to 72 hours^21^, the US Centers for Disease Control and Prevention suggest a system in which each healthcare worker is allocated five respirators for daily use in strict rotation, using careful storage to essentially ‘quarantine’ the devices. The US Centers for Disease Control and Prevention suggest that if five respirators are not available for each worker, then decontamination may be necessary, potentially suggesting that this should be an initial strategy prior to attempting reprocessing. We found no systematic review evidence testing this approach, and neither Public Health England or European Centre for Disease Prevention and Control explicitly discuss re-use with or without reprocessing. Moreover, data from the study cited showed that the 72 hour period was just the length of the measurement period, and that survival followed a decay curve, not a fixed “all-dead” time, implying that the length of time an amount greater than or equal to the infective dose survives depends on the starting quantity of virions^21^.

The systematic reviews included laboratory studies to test whether reprocessing changed the properties of a respirator. None of the reviews or research reported in guidance documents described practical or operational studies of facilitating re-use or reprocessing of respirators at scale. This is a key gap, given existing strain on healthcare resources and the requirement to ensure that a respirator remains matched to a healthcare worker throughout its use.

## Discussion

### Statement of principal findings

There are five key findings from this review. First, while extended use or re-use of single-use surgical masks or respirators (with or without reprocessing) is generally not recommended, guidance from various organisations supports such measures (preferably extended use rather than re-use) as a last-resort measure during critical shortage. Second, comparisons across guidance documents and systematic reviews highlight limited evidence, varying levels of detail, and areas of inconsistency, especially in relation to reuse of respirators (with or without reprocessing) during and after aerosol generating procedures. Third, the reprocessing of surgical masks is not recommended. Fourth, reprocessing of respirators under controlled and standardised conditions is recommended, but there is inconsistency regarding how or when this should take place. Fifth, where extended use or re-use is being practised, healthcare facilities and institutions should ensure that policies and systems are in place to enable these practices to be carried out in the safest way possible in line with available guidance.

### Strengths and limitations

This review was conducted in adherence with current Cochrane rapid review guidance and used more than one author for searching, screening, data extraction and quality appraisal of included studies. The interdisciplinary nature of the research team was a particular strength and included frontline healthcare workers (XHC, LR) and individuals with expertise in occupational medicine (AA), infection control (XHC, LR), respiratory protective equipment design and performance (SS, IT), emergency nursing (YC), evidence synthesis (ET, YC, DD, TG, CB, ST) and an information specialist (MS). Owing to the need for timely review production, we limited our search for guidance to four major international health organisations and we did not search exhaustively for primary studies. At the time of writing, the included systematic reviews are non-peer reviewed pre-prints, but this reflects the rapid emergence of new evidence in this field. Recent research has compared international regulations regarding re-use and extended use of filtering facepiece respirators^6^, and guidance on respiratory protective equipment more broadly^22^. However, the former did not address surgical masks, and neither study compared recommendations with current evidence. Our study synthesises and integrates recommendations and evidence for single-use surgical masks and respirators to enable a clearer understanding of current evidence, gaps and facilitate evidence-informed decision-making in this area.

### Meaning of the study: implications for clinicians and policymakers

This study has emphasised the numerous risks relating to the extended use, re-use or reprocessing of single-use surgical masks and respirators. Guidance is unanimous that these practices should be considered only in situations of critical shortage, after all other strategies have been employed to minimise strain on supply.

Where extended use or re-use is unavoidable, risks should be carefully assessed and policies and decisionmaking should be made on best available evidence. Where evidence is lacking or unclear, difficult judgements need to be made to balance current safety of staff over conservation of supply with a view to future protection of staff. Given the rapidly growing research literature on this, guidance should be regularly reviewed and updated.

Surgical masks and respirators have different properties and functions. They should be distinguished clearly in policies and guidance, which should also take into consideration the variability of respirator models and manufacturer guidance. Policies should also be developed that address re-use clearly in relation to different reprocessing methods, and also address use and re-use of respirators in different situations e.g. during and after aerosol generating procedures. Policies on re-use and extended use need to address both individual factors (e.g. regarding discarding, safe use and duration, and number of uses), and organisational factors (such as management of the supply chain, safe reprocessing and storage, staff training, and monitoring and evaluation of practice).

Certain steps can be taken to mitigate risk. This includes extending use of single-use masks or respirators before resorting to re-use and regularly inspecting masks and respirators for integrity, visible damage and fit. Knowing when equipment must be discarded is crucial, as is appropriate storage and clear labelling of respirators between use, to avoid cross-use between workers. Organisations should ensure that adequate training in this is provided.

Policy guidance emphasises the need to assess the contagion risk of an encounter and use the recommended protective ensemble for that situation^5^. Surgical masks and filtering facepiece respirators are only one component of personal protective equipment, which typically includes gloves, long-sleeved fluid repellent gown, and eye protection^2^. Safe donning and doffing are critical^2^. Personal protective equipment is considered as one of the last lines of defence within the hierarchy of infection control measures which also include administrative and environmental and engineering controls.

### Unanswered questions and future research

There are several areas warranting further investigation in relation to reprocessing. In the current context, research is needed which explores the impact of respirator decontamination methods on SARS-CoV-2, taking heterogeneity of models into account. There is also a paucity of research exploring the impact of decontamination on important outcomes such as respirator fit, user comfort and safety and the feasibility of reprocessing methods at scale.

Despite consideration in some guidance documents for the extended use and re-use of surgical masks in crisis capacity situations, there is limited evidence in this area. There is a need for further research regarding the effects of extended use of masks and respirators on outcomes such as fit and user comfort to determine the number of uses possible, and the optimum length of extended use. Our recent review of respirator performance and standards found that all respirator types carry a burden to the user of discomfort and interference with communication^23^. Houghton et al. recently identified user comfort as an important factor influencing adherence with infection prevention and control guidelines^24^; this becomes particularly important when worn for extended periods.

### Conclusion

Extended use and re-use of single-use surgical masks and respirators (with or without reprocessing) should only be considered in situations of critical shortage. Where extended use or re-use is being practised, healthcare organisations should ensure that policies and systems are in place to ensure these practices are carried out as safely as possible and in line with available guidance. Areas of guidance lacking clarity and consistency warrant further attention and investigation.

## Data Availability

This is a systematic review with narrative synthesis therefore the data associated is minimal. All supplementary files are provided. Protocols and raw data (e.g. data extraction files) are available on OSF at https://osf.io/7c6rs/

## Contributorship and guarantor

The article was collaboratively developed as part of a wider series of evidence reviews on personal protective equipment edited by TG and overseen by the Oxford Covid-19 Evidence Review Service. TG, IT, SS, AA and members of the Oxford Covid-19 Evidence Review Service conceptualised the review. ET led the design and execution of the study to align with formal systematic review guidance. MS, ET, IT and ST undertook searches and screening. ET, YC and DD conducted data extraction. ET led the synthesis with input from YC and CB. XHC and LR provided specialist infection control expertise. SS and IT provided technical expertise on masks and respirators. ET, CB and YC wrote the first draft of the paper, to which all authors made contributions. All authors approved the final manuscript. ET is corresponding author and guarantor.

## Acknowledgements

The authors thank Kamlesh Khunti, Sebastian Straube, Quentin-Durand Moreau and Tanya Jackson of the Oxford Covid-19 Evidence Review Service team for their contribution to the review conceptualisation, and Vicki Martin for her contribution to graphic design.

## How patients were involved in the creation of this article

Members of the public were not involved in the review or the writing of the paper.

## References

1. Adhanom Ghebreyesus T. WHO Director-General’s opening remarks at the media briefing on COVID19. https://www.who.int/dg/speeches/detail/who-director-general-s-opening-remarks-at-the-media-briefing-on-covid-19---11-may-2020, 2020.

2. Verbeek JH, Rajamaki B, Ijaz S, et al. Personal protective equipment for preventing highly infectious diseases due to exposure to contaminated body fluids in healthcare staff. Cochrane Database of Systematic Reviews 2020(4) doi: 10.1002/14651858.CD011621.pub4

3. World Health Organisation. Shortage of personal protective equipment endangering health workers worldwide. Geneva: World Health Organisation, 2020.

4. US Centers for Disease Control and Prevention. Recommended guidance for extended use and limited reuse of n95 filtering facepiece respirators in healthcare settings-NIOSH Workplace Safety and Health Topic 2020 [updated March 27, 2020. Available from: https://www.cdc.gov/niosh/topics/hcwcontrols/recommendedguidanceextuse.html.

5. World Health Organization. Rational use of personal protective equipment for Coronavirus disease (COVID-19) and considerations during severe shortages: interim guidance, 6 April 2020: World Health Organization; 2020 [Available from: https://www.who.int/publications-detail/rational-use-of-personal-protective-equipment-for-coronavirus-disease-(covid-19)-and-considerations-during-severe-shortages.

6. Kobayashi LM, Marins BR, Costa PCdS, et al. Extended use or reuse of N95 respirators during COVID-19 pandemic: an overview of national regulatory authorities’ recommendations. Infect Control Hosp Epidemiol 2020:1–43. doi: 10.1017/ice.2020.173 [published Online First: 2020/04/22]

7. Garritty C, Gartlehner G, Kamel C, et al. Cochrane Rapid Reviews Interim Guidance from the Cochrane Rapid Reviews Methods Group 2020, 2020.

8. Toomey E. Overview of recommendations and evidence for reuse and/or extended use of respiratory protective equipment (RPE) for the prevention of COVID-19: Protocol. Open Science Framework, 2020.

9. Moher D, Liberati A, Tetzlaff J, et al. Preferred Reporting Items for Systematic Reviews and Meta-Analyses: The PRISMA Statement. PLOS Medicine 2009;6(7):e1000097. doi: 10.1371/journal.pmed.1000097

10. US Centers for Disease Control and Prevention. Strategies for optimizing the supply of facemasks: CDC Atlanta, GA; 2020 [Available from: https://www.cdc.gov/coronavirus/2019-ncov/hcp/ppe-strategv/face-masks.html.

11. US Centers for Disease Control and Prevention. Decontamination and reuse of filtering facepiece respirators 2020 [Available from: https://www.cdc.gov/coronavirus/2019-ncov/hcp/ppe-strategy/decontamination-reuse-respirators.html9.

12. European Centre for Disease Prevention and Control. Cloth masks and mask sterilisation as options in case of shortage of surgical masks and respirators 2020 [Available from: https://www.ecdc.europa.eu/sites/default/files/documents/Cloth-face-masks-in-case-shortage-surgical-masks-respirators2020-03-26.pdf.

13. Public Health England. Considerations for acute personal protective equipment (PPE) shortages 2020 [updated April 27, 2020. Available from: https://www.gov.uk/government/publications/wuhan-novel-coronavirus-infection-prevention-and-control/managing-shortages-in-personal-protective-equipment-ppe.

14. Gertsman S, Agarwal A, O’Hearn K, et al. Microwave-and Heat-Based Decontamination of N95 Filtering Facepiece Respirators (FFR): A Systematic Review. 2020

15. O’Hearn K, Gertsman S, Sampson M, et al. Decontaminating N95 masks with Ultraviolet Germicidal Irradiation (UVGI) does not impair mask efficacy and safety: A Systematic Review. 2020

16. O’Hearn K, Gertsman S, Webster R, et al. Efficacy and Safety of Disinfectants for Decontamination of N95 and SN95 Filtering Facepiece Respirators: A Systematic Review. 2020

17. Fischer R, Morris DH, van Doremalen N, et al. Assessment of N95 respirator decontamination and re-use for SARS-CoV-2. medRxiv 2020:2020.04.11.20062018. doi: 10.1101/2020.04.11.20062018

18. Kumar A, Kasloff SB, Leung A, et al. N95 Mask Decontamination using Standard Hospital Sterilization Technologies. medRxiv 2020:2020.04.05.20049346. doi: 10.1101/2020.04.05.20049346

19. Radonovich LJ, Cheng J, Shenal BV, et al. Respirator Tolerance in Health Care Workers. JAMA 2009;301(1):36–38. doi: 10.1001/jama.2008.894

20. Radonovich LJ, Yanke R, Cheng J, et al. Diminished Speech Intelligibility Associated with Certain Types of Respirators Worn by Healthcare Workers. J Occup Environ Hyg 2009;7(1):63–70. doi: 10.1080/15459620903404803

21. van Doremalen N, Bushmaker T, Morris D, et al. Aerosol and surface stability of HCoV-19 (SARS-CoV-2) compared to SARS-CoV-1. medRxiv 2020:2020.03.09.20033217. doi: 10.1101/2020.03.09.20033217

22. Birgand G, Mutters NT, Otter J, et al. Analysis of national and international guidelines on respiratory protection equipment for COVID-19 in healthcare settings. medRxiv 2020:2020.04.23.20077230. doi: 10.1101/2020.04.23.20077230

23. Burton C, Coles B, Anisesh A, et al. Performance and impact of disposable and reusable respirators for healthcare workers during pandemic respiratory disease: a rapid evidence review. medRxiv 2020:2020.05.21.20108233. doi: 10.1101/2020.05.21.20108233

24. Houghton C, Meskell P, Delaney H, et al. Barriers and facilitators to healthcare workers’ adherence with infection prevention and control (IPC) guidelines for respiratory infectious diseases: a rapid qualitative evidence synthesis. Cochrane Database of Systematic Reviews 2020(4) doi: 10.1002/14651858.CD013582

